# Interference between Venus-A Valve and Anterior Mitral Valve Leaflets after Transcatheter Aortic Valve Replacement: Insight from FEops HEARTguide simulation

**DOI:** 10.1101/2023.04.14.23288608

**Authors:** Yong Wang, Ting Liu, Ying Zeng, Nic Debusschere, Giorgia Rocatello, Sihang Cheng, Ping Li, Dehui Qian, Shiyong Yu, Jun Jin

**Affiliations:** Department of Cardiology, Institute of Cardiovascular Research, The Second Affiliated Hospital of Army Medical University, Chongqing, China; FEops NV, Ghent, Belgium; Venus Medtech, Hangzhou, China

**Keywords:** transcatheter aortic valve replacement, anterior mitral leaflet, interference, computer simulation

## Abstract

**Background:** Scarce data exist regarding the occurrence of mitral valve interference after transcatheter aortic valve replacement (TAVR) with Venus-A valve implantation. Several case reports have noted that the anterior mitral leaflet (AML) is mechanically affected by the prosthesis frame, particularly when implanted in a low position. This study aimed to investigate the potential factors influencing the clinical outcomes of AML interference after Venus-A valve implantation.

**Methods:** We retrospectively included 20 severe aortic valve stenosis patients who had undergone TAVR and had been implanted with the Venus-A valve at our hospital between October 2020 and June 2021. Pre- and post-procedural CT scans were used for the FEops HEARTguide simulation. Anatomically influencing factors were measured using the 3mensio software and derived from the FEops HEARTguide. The prosthesis-AML interference (PAI) was defined when it met both of two criteria:1) significant interference and limited AML movement shown by transthoracic or transoesophageal echocardiography, and 2) more than half cell intersection between the simulated Venus-A valve and the reconstructed AML revealed by the FEops HEARTguide. Anatomical factors and clinical outcomes were compared between the PAI and non-PAI groups.

**Results:** Nine PAI patients and 11 non-PAI cases were identified. PAI was associated with shorter mitral-aortic annulus distance (2.7±1.7 mm vs 5.0±2.2 mm, *P* = 0.019), larger prosthesis valve size (*P* = 0.013), deeper implantation (12.2±3.3 mm vs 6.2±2.9 mm at non-coronary cusp side, *P* < 0.001) and less calcification of non-coronary cusp (median calcification score, 52.2 mm^3^ vs 156.0 mm^3^, *P* = 0.046). Regarding the clinical impact, PAI was associated with a higher rate of moderate or severe perivalvular leakage before discharge than those associated with the absence of PAI, with no difference in haemodynamic parameters and incidence of adverse events at the 30-day and 12-month follow-ups between the groups.

**Conclusions:** Interference between the Venus-A prosthesis valve and AML after TAVR was associated with a shorter mitral-aortic annulus distance, larger prosthesis usage, greater implantation depth, and less calcification of the non-coronary cusp. However, further studies are required to explore its long-term clinical impact.

## 1 Introduction

Transcatheter aortic valve replacement (TAVR) is the leading therapeutic strategy for aortic valve replacement in patients with severe symptomatic AS (1). As the anterior mitral annulus is anatomically linked to both the left and non-coronary aortic cusps through a shared fibrous rim (2), the prosthesis frame mechanically interferes with the anterior mitral leaflet (AML), especially when implanted in a low position. Several case reports have reported that deep aortic prosthetic valve implantation may impair adequate AML opening (3,4), even led to direct erosive perforation and infective endocarditis (5-7). However, the existing knowledge about this rarely described complication remains limited.

A self-expandable valve purportedly has a higher risk of AML interference than that associated with a balloon-expandable valve owing to its long prosthesis frame [4]. The self-expandable Venus-A valve was the first approved transcatheter heart prosthesis and is the most widely used valve in Mainland China. The design characteristics of the Venus-A valves have been previously reported in detail (8). Data on mitral valve interference after Venus-A valve implantation are scarce. Patient-specific computational modelling of TAVR with the FEops HEARTguide (FEops nv, Ghent, Belgium) based on pre-procedural dual-source computed tomography (DSCT) could accurately predict device-anatomy interactions between a self-expandable transcatheter device model and the surrounding anatomical structures (9-11), in both tricuspid and bicuspid aortic valve anatomy (11-14). Three-dimensional computer models can provide detailed insights to help investigate the interference between the AML and the prosthesis frame.

In this exploratory study, we sought to explore prosthesis-AML interference in patients with aortic valve stenosis treated with the Venus-A valve through patient-specific computer simulation.

## 2 Materials and Methods

### 2.1 Patient population

This single-centre, retrospective, observational study included 20 patients with severe aortic stenosis who successfully underwent TAVR at our hospital. All patients who had undergone transfemoral TAVR using a first-generation self-expandable Venus-A valve (Venus MedTech Inc., Hangzhou, China), and those who had undergone a second valve implantation (valve-in-valve) were excluded. The design characteristics of the Venus-A valves have been previously reported in detail (8). Preprocedural and postprocedural computed tomography scans were performed in 20 and 17 patients, respectively, using DSCT (SOMATOM Definition Flash, Siemens Medical Solutions, Germany). The heart team discussed the indications for TAVR, and the size of the prostheses was determined based on the aortic root DSCT.

This study was approved by the Research Ethics Committee of the Second Affiliated Hospital (Xinqiao Hospital) of the Army Military Medical University, and the requirement for informed consent was waived because of its retrospective design.

### 2.2 Data collection

Baseline clinical information, echocardiographic, DSCT, procedural, and clinical follow-up data were collected. All patients underwent echocardiography and electrocardiography before discharge and at both 30-day and 12-month follow-ups. Postprocedural DSCT was performed 6– 12 months after TAVR (mean 8.2 months). DSCT data were retrospectively analysed using the 3mensio software (Pie Medical, Bilthoven, Netherlands). The aortic root structure was measured in the 40% systolic phase. The aortic annulus was defined as the virtual basal plane containing the basal attachment of the three aortic cusps. Sizes of the annulus, left ventricular outflow tract, sinotubular junction, and ascending aorta were measured. Aortic valve morphology was recorded using the Sievers classification (15). The aortic valve calcification volume was automatically measured using a calcification threshold of 850 HU. Clinical events were recorded according to Valve Academic Research Consortium-3 (VARC-3) criteria (16). Two experienced sonographers independently checked the position of the TAVR prosthesis and its relationship with the AML, and evaluated the morphology and motion of the AML. The degree of their interaction was graded as follows:0, no interference between prosthesis and AML; 1, interference but the motion of AML was not obviously affected; and 2, significant interference and limited AML movement. The kappa coefficient of agreement was 0.92. Prosthesis AML interference (PAI) was defined if met both of the 2 criteria:1) significant interference and limited AML movement shown by transthoracic or transoesophageal echocardiography, and 2) more than half-cell intersection between the simulated Venus-A valve and the reconstructed AML revealed by computer simulation.

### 2.3 Computer simulation

Briefly, preprocedural and postprocedural CT scans were obtained from 20 and 17 patients, respectively. Preprocedural cardiac CT images were used to reconstruct finite element models of the aortic root, including native calcified aortic leaflets, left ventricle, left atrium, and mitral leaflets **(Figure 1a-1b-1c)**. Different material properties were used to model the native aortic wall (E=0.6 MPa, ν=0.3), native leaflet tissue (E=2 MPa, ν=0.45) and calcium nodules (E=4 MPa, ν=0.3, Yield stress = 0.6 MPa) (9,10). The left ventricle, left atrium, and mitral leaflets were used for visualisation purposes only to evaluate the interference between the AML and simulated device. Therefore, no material properties are assigned to these structures.

**Figure 1.**
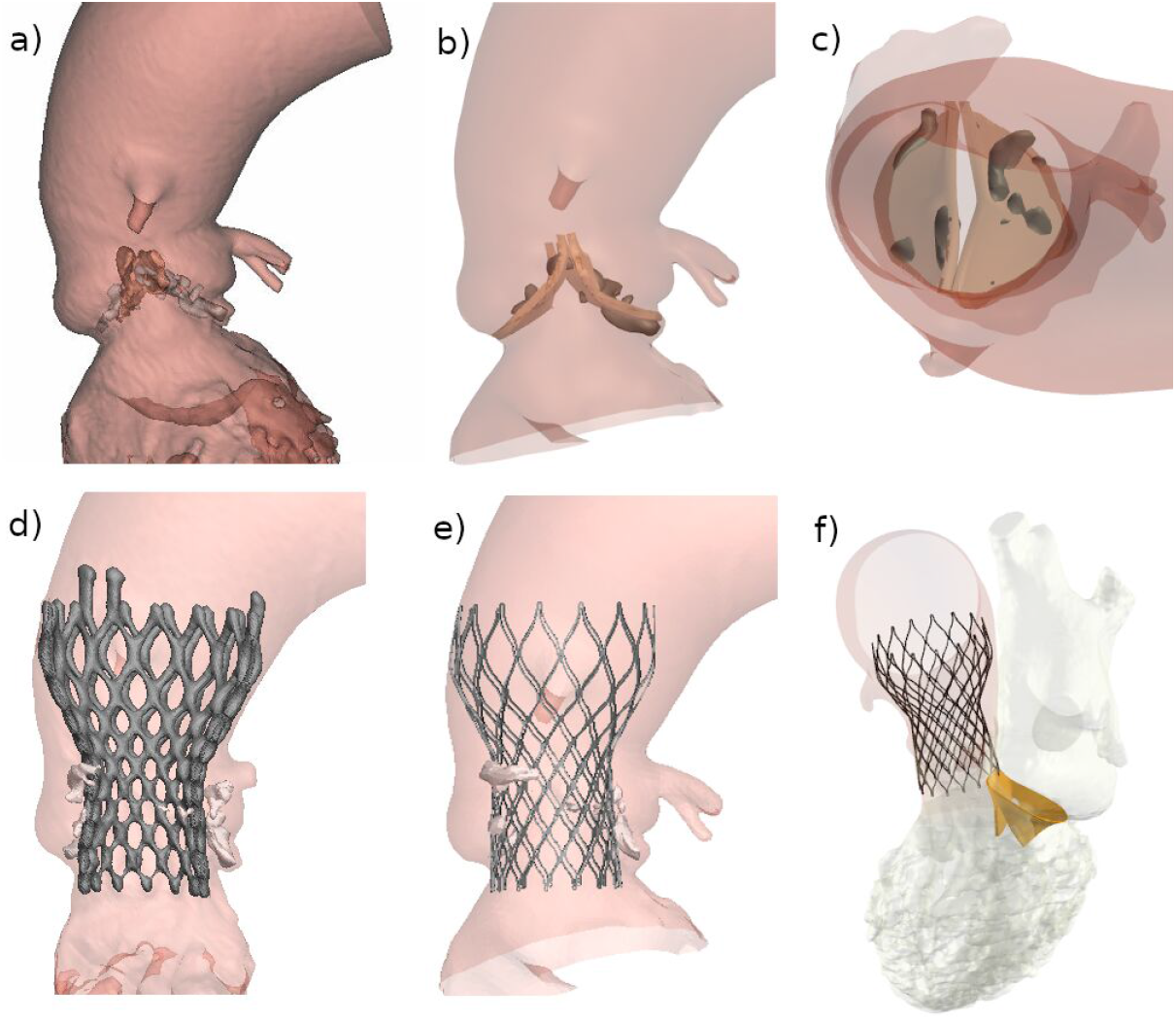
Patient-specific computer simulation workflow. a) Aortic root anatomical structures including native calcified leaflets segmented from preoperative CT images in Mimics (Materialize, Leuven); b,c) Aortic root anatomical structures reconstructed with FEops HEARTguide from frontal and top view respectively; d) segmentation of the VenusA valve based on postoperative CT images in Mimics; e) simulation of the VenusA implantation using FEops HEARTguide; f) evaluation of the interaction between the simulated Venus-A valve and the AML reconstructed with FEops HEARTguide.

In the finite element analysis simulation, the crimped Venus-A valve model was positioned coaxially within the aortic root and deployed by retracting the sheath. In each simulation, the device size and position were consistent with those used in the clinical procedure. Device implantation was iteratively simulated until the final device position matched the actual depth of implantation, as measured from post-procedural CT images in 17 patients (**Figure 1d-1f**). In 3 patients, the final implantation depth at the noncoronary and left coronary cusps was derived from DSA.

### 2.4 Morphological Interference Analysis

The inner linings of the left ventricle and left atrium were segmented from the CT images. The mitral and aortic annuli were manually indicated on CT images. The mitral valve leaflets were reconstructed by mapping a template mesh with the leaflets visible on CT images. The left ventricle, left atrium, and mitral leaflets were used for visualisation purposes only, whereas the aortic annual plane and mitral annulus were used to calculate the mitral-aortic annulus length (distance) and angle.

A reference plane was assigned **to each mitral valve (Figure 2)**, with its origin at the centre of the mitral annulus and its normal vector aligned with the line connecting the posteromedial and anterolateral trigones. This plane intersects the mitral and aortic annuli. The mitral-aortic annulus length was defined as the minimum distance between the intersection points. The mitral and aortic annulus normal vectors were defined by determining the best-fitting planes using both closed curves. The mitral-aortic annulus angle was defined as the angle between the respective normal vectors. Morphological interference was defined as more than half of the cells of the Venus A valve intersecting the reconstructed AML in the simulation results. **Figure 3** shows two cases with no morphological interference **(Figure 3a-3b)** and one with morphological interference **(Figure 3c). Supplementary Figure 1** defined “half-cell intersection”.

**Figure 2.**
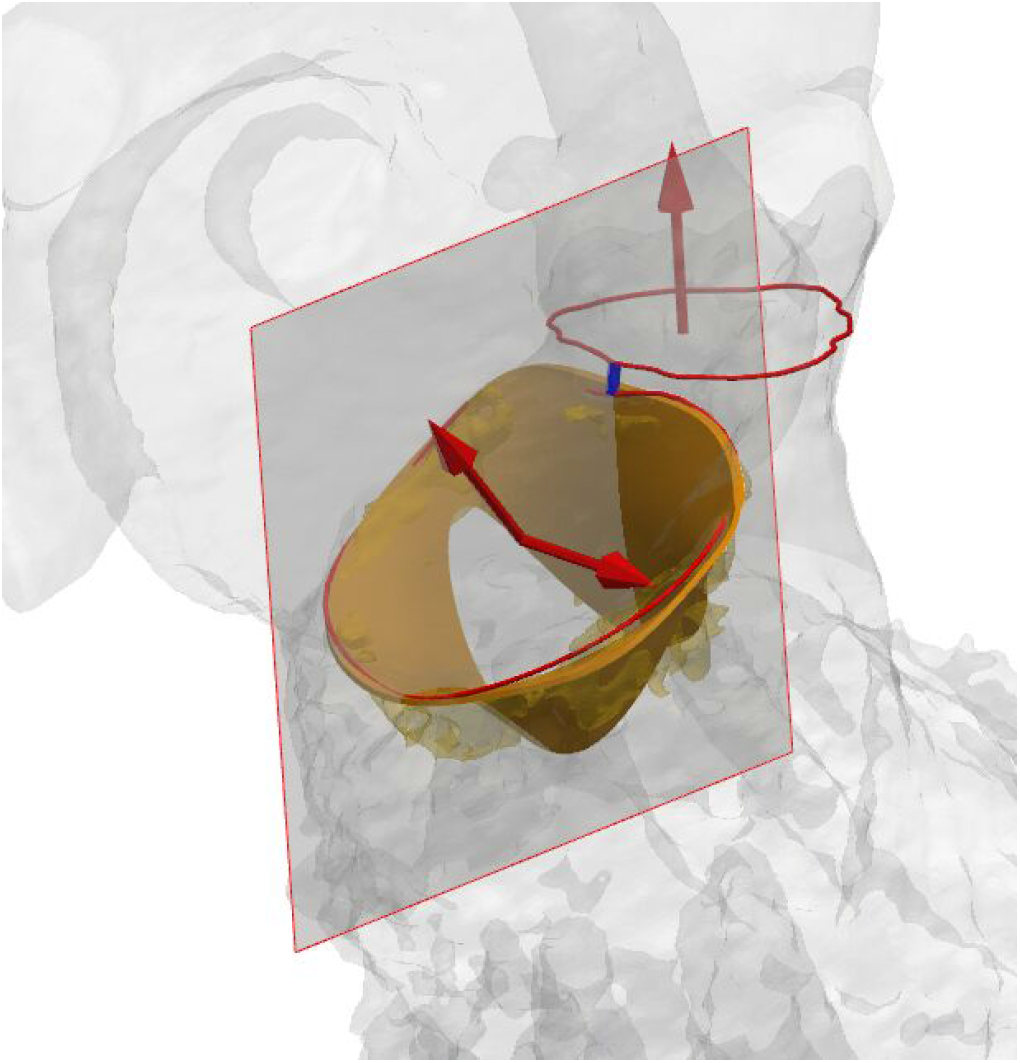
Definition of mitral-aortic annulus distance (blue line) and mitral-aortic annulus angle. Shown is a reference plane through the center of the mitral annulus with normal vector aligned with the postero-medial and antero-lateral trigones, and the best fitting normal vectors of the mitral annulus and aortic annulus.

**Figure 3.**
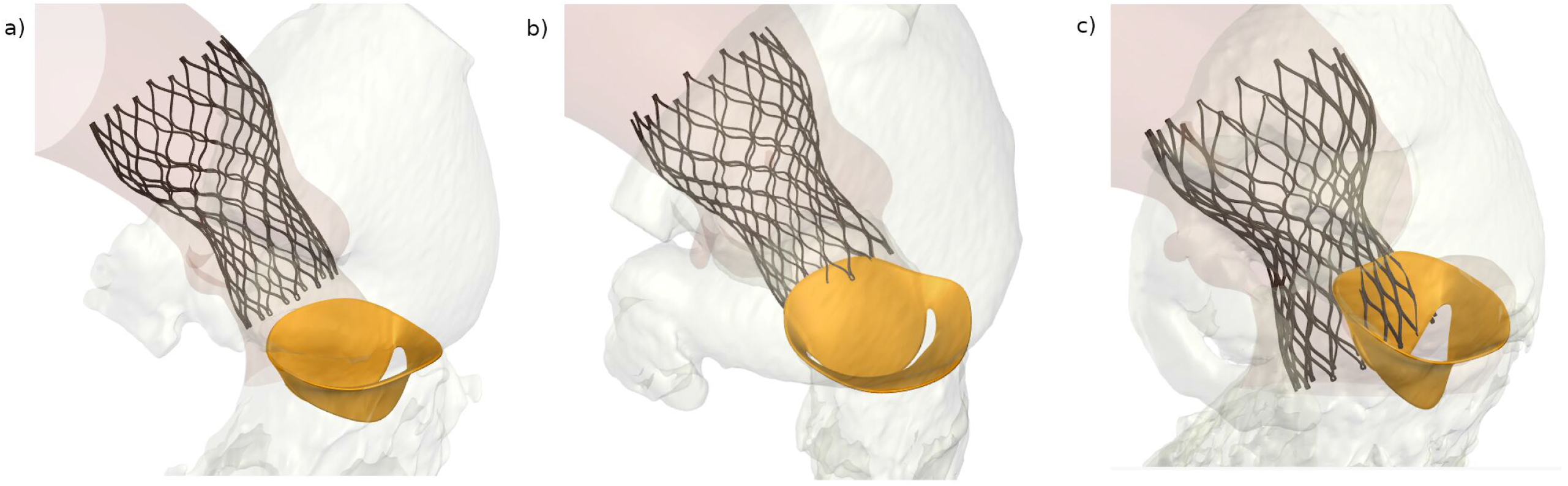
Representative cases of PAI and non-PAI. a) Case with no intersection between the simulated Venus-A valve and the reconstructed AML resulting in no morphological interference; b) case with limited intersection between the simulated Venus-A valve and the reconstructed AML (intersection <half cell) resulting in no morphological interference; c) case with large intersection between the simulated Venus-A valve and the reconstructed AML (intersection >half cell) resulting in morphological interference.

### 2.5 Statistical Analysis

Continuous variables with normal distribution are expressed as mean ± standard deviation; those with skewed distribution are expressed as median (lower and upper quartile), while categorical variables are reported as numbers (proportion). The independent sample *t*-test or Mann-Whitney *U* test was used to compare the means between the two groups, and Fisher’s exact test was used for categorical variables. All statistical tests were 2-tailed, with *P* <0.05. considered statistically significant. Statistical analyses were performed using SPSS (version 26.0; Chicago, Armonk, NY, USA).

## 3 Results

As presented by the FEops HEARTguide, six cases were identified without an intersection between the prosthesis frame and AML **(Figure 3a)**, five had an intersection < half cell **(Figure 3b)**, and nine cases were identified with more than half of the cells **(Figure 3c)**. Consistently, echocardiography showed that all 9 patients with an intersection of more than half of the cells showed significant interference and limited AML movement, whereas those with an intersection <half-cell showed no limited AML movement. Therefore, 9 PAI cases and 11 non-PAI cases were identified.

As shown in **Table 1**, the groups did not differ in terms of sex, age, and other clinical characteristics, while the presence of atrial fibrillation (*P* = 0.050) and more than mild mitral regurgitation was higher (*P* = 0.028) in the PAI group. The left ventricular anteroposterior diameter was larger (*P* = 0.028), while the ejection fraction and fractional shortening were lower in the PAI group (both *P* <0.05).

**Table 1.** Baseline characteristics of PAI group and Non-PAI group Data are presented as mean ± standard deviation or median (lower quartile, upper quartile). PAI, Prosthesis-anterior mitral leaflet interference; NYHA, New York Heart Association; STS, Society of Thoracic Surgeons. NCC, Non coronary cusp; RCC, Right coronary cusp; LCC, Left coronary cusp.

|  | PAI group (n=9) | Non-PAI group (n=11) | P value |
| --- | --- | --- | --- |
| **Clinical data** |  |  |  |
| Male gender | 3/9 | 5/11 | 0.670 |
| Age (yrs) | 69.1±6.2 | 72.3±7.1 | 0.302 |
| Body mass index (Kg/m<sup>2</sup>) | 24.5±4.8 | 25.4±4.3 | 0.645 |
| Coronary heart disease | 2/9 | 2/11 | 1.000 |
| Atrial fibrillation | 5/9 | 1/11 | 0.050* |
| Hypertension | 4/9 | 5/11 | 1.000 |
| Diabetes | 0/9 | 2/11 | 0.479 |
| NYHA class III/IV | 7/9 | 5/11 | 0.197 |
| STS score (%) | 4.5±1.6 | 4.2±1.8 | 0.726 |
| **Ultrasound data** |  |  |  |
| Left atrium anteroposterior diameter (mm) | 41.4±6.9 | 37.1±4.9 | 0.115 |
| Left ventricle anteroposterior diameter (mm) | 52.0±6.0 | 45.8±4.2 | 0.015* |
| Interventricular septum thickness (mm) | 12.8±1.4 | 13.7±2.2 | 0.283 |
| Ejection fraction (%) | 59.9±6.0 | 64.8±3.5 | 0.039* |
| Fraction shortening (%) | 31.9±4.0 | 35.4±2.6 | 0.036* |
| Peak aortic flow velocity (cm/s) | 466.9±58.1 | 507.6±70.5 | 0.182 |
| Mean aortic valve pressure gradient (mmHg) | 49.0±12.9 | 57.6±16.0 | 0.207 |
| ≥Mild mitral regurgitation | 8/9 | 4/11 | 0.028* |
| ≥Moderate mitral regurgitation | 3/9 | 0/11 | 0.074 |
| ≥Moderate aortic regurgitation | 4/9 | 2/11 | 0.336 |
| **Feops data** |  |  |  |
| Distance between aortic / mitral planes (mm) | 2.7±1.7 | 5.0±2.2 | 0.019* |
| Angle between aortic / mitral planes (degree) | 131.4±11.6 | 126.2±11.5 | 0.333 |
| **CT data** |  |  |  |
| Bicuspid | 5/9 | 7/11 | 1.000 |
| Type 0 | 4/5 | 3/7 |  |
| Type 1 | 1/5 | 4/7 |  |
| **Aortic annulus** |  |  |  |
| Mean diameter (mm) | 24.2±1.6 | 22.2±1.6 | 0.016* |
| Perimeter (mm) | 76.5±5.6 | 69.5±5.4 | 0.011* |
| Area (mm<sup>2</sup>) | 451.6±57.5 | 382.3±54.0 | 0.013* |
| Left ventricular outflow tract perimeter (mm) | 83.9±7.5 | 64.8±15.4 | 0.003* |
| Sinotubular Junction mean diameter (mm) | 31.8±4.6 | 29.0±3.3 | 0.144 |
| Aortic angulation (degree) | 49.7± 8.7 | 50.5±15.8 | 0.895 |
| NCC calcification score (mm<sup>3</sup>) | 52.2 (22.9, 79.1) | 156.0 (3.1, 246.2) | 0.046* |
| RCC calcification score (mm<sup>3</sup>) | 129.6 (51.7, 254.6) | 167.5 (85.6, 268.3) | 0.412 |
| LCC calcification score (mm<sup>3</sup>) | 115.7 (37.2, 290.5) | 43.5 (4.9, 146.3) | 0.295 |
| Total leaflet calcification score (mm<sup>3</sup>) | 243.6 (127.3, 589.1) | 397.3 (144.0, 588.6) | 0.656 |

The anatomical characteristics of the patients are shown in **Table 2**. According to the computer simulation parameters, the length between aortic-mitral annulus was shorter in PAI cases (2.7±1.7 mm vs 5.0±2.2 mm, *P* = 0.019), but the angle showed no difference **(Figure 2)**. According to the CT parameters, PAI was associated with a larger annulus and LVOT (all *P* <0.05). Nevertheless, less calcification of the non-coronary cusp was observed in the PAI group (*P* = 0.046). The procedural characteristics and in-hospital clinical outcomes are listed in **Table 3**. The proportion of larger size prosthesis application was higher (*P* = 0.013), and the implantation depth was significantly deeper in patients with PAI (12.2±3.3 mm vs 6.2±2.9 mm at NCC side, *P* < 0.001, and 14.3±4.7 mm vs 7.7±3.1 mm at LCC side, *P* = 0.002, respectively). Moreover, the incidence of moderate perivalvular leakage (PVL) was higher in the PAI group (5/9 vs 0/11, *P* = 0.008). The incidence of new-onset left bundle branch block was higher in the PAI group; however, the difference was not statistically significant (*P* = 0.070).

**Table 2.** Intra-Procedural data and in-hospital outcomes of the two groups Data are presented as mean ± standard deviation or median (lower quartile, upper quartile). PAI, Prosthesis-anterior mitral leaflet interference; NYHA, New York Heart Association; STS, Society of Thoracic Surgeons. NCC, Non coronary cusp; LCC, Left coronary cusp. LBBB, left bundle branch block. NA, Not Applicable.

|  | PAI group (n=9) | Non-PAI group (n=11) | *P value* |
| --- | --- | --- | --- |
| *Prosthesis size* |  |  | **0.013\*** |
| *L23* | 2/9 | 9/11 |  |
| *L26* | 4/9 | 2/11 |  |
| *L29* | 3/9 | 0/11 |  |
| Implantation depth at NCC (mm) | 12.2±3.3 | 6.2±2.9 | **<0.001\*** |
| Implantation depth at LCC (mm) | 14.3±4.7 | 7.7±3.1 | **0.002\*** |
| Pre dilation | 9/9 | 11/11 | NA |
| Post dilation | 1/9 | 6/11 | 0.070 |
| Coronary obstruction | 0/9 | 1/11 | 1.000 |
| New onset atrial fibrillation | 0/9 | 1/11 | 1.000 |
| New onset LBBB | 7/9 | 3/11 | 0.070 |
| Need of permanent pacemaker | 1/9 | 2/11 | 1.000 |
| Major bleeding | 0/9 | 1/11 | 1.000 |
| ≥Moderate perivalvular leakage | 5/9 | 0/11 | **0.008\*** |
| Peak aortic flow velocity (cm/s) | 271.7±33.2 | 273.5±50.5 | 0.928 |
| Mean aortic valve pressure gradient (mmHg) | 15.0±3.8 | 16.9±7.5 | 0.497 |
| ≥Mild mitral regurgitation | 7/9 | 3/11 | 0.070 |
| ≥Moderate mitral regurgitation | 2/9 | 1/11 | 0.566 |

Data are presented as mean ± standard deviation.
PAI, Prosthesis-anterior mitral leaflet interference; NCC, Non coronary cusp; LCC, Left coronary cusp. LBBB, left bundle branch block. NA, Not Applicable.

**Table 3.** The 30-day and 12-month clinical outcomes of the two groups Data are presented as mean ± standard deviation. PAI, Prosthesis-anterior mitral leaflet interference; NYHA, New York Heart Association; NA, Not Applicable.

|  | PAI group (n=9) | Non-PAI group (n=11) | *P value* |
| --- | --- | --- | --- |
| **30-day outcome** |  |  |  |
| Peak aortic flow velocity (cm/s) | 256.5±42.3 | 281.7±60.1 | 0.483 |
| Mean aortic valve pressure gradient (mmHg) | 13.6±4.5 | 17.5±7.6 | 0.312 |
| Mean mitral valve pressure gradient (mmHg) | 1.6±0.6 | 1.5 (1.0, 2.1) | 0.945 |
| ≥Mild mitral stenosis | 0/9 | 0/11 | NA |
| ≥Moderate mitral regurgitation | 2/9 | 2/11 | 0.625 |
| ≥Moderate perivalvular leakage | 4/9 | 1/11 | 0.127 |
| NYHA class III/IV | 4/9 | 1/11 | 0.127 |
| Stroke | 0/9 | 0/11 | NA |
| All-cause mortality | 0/9 | 0/11 | NA |
| Infective endocarditis | 0/9 | 0/11 | NA |
| Mitral valve perforation | 0/9 | 0/11 | NA |
| **12-month outcome** |  |  |  |
| Peak aortic flow velocity (cm/s) | 279.7±22.6 | 292.6±64.1 | 0.681 |
| Mean aortic valve pressure gradient (mmHg) | 15.0±2.9 | 19.4±8.2 | 0.191 |
| Mean mitral valve pressure gradient (mmHg) | 1.9±1.4 | 1.5 (1.0, 2.0) | 0.964 |
| ≥Mild mitral stenosis | 0/9 | 0/11 | NA |
| ≥Moderate mitral regurgitation | 3/9 | 1/11 | 0.217 |
| ≥Moderate perivalvular leakage | 4/9 | 1/11 | 0.127 |
| NYHA class III/IV | 1/9 | 0/11 | 0.450 |
| Stroke | 0/9 | 0/11 | NA |
| All-cause mortality | 0/9 | 0/11 | NA |
| Infective endocarditis | 0/9 | 0/11 | NA |
| Mitral valve perforation | 0/9 | 0/11 | NA |

Data are presented as mean ± standard deviation or median (lower quartile, upper quartile).

Regarding clinical outcomes, no significant differences were observed in aortic flow velocity, aortic pressure gradient, or clinical events such as mortality, stroke, PVL, and heart failure between the two groups at both the 30-day and 12-month follow-ups. In particular, we observed special adverse consequences of PAI according to previous reports (3-6), and no new-onset mitral stenosis, infective endocarditis, or mitral valve perforation occurred in either group (**Table 3)**.

## 4 Discussion

To our knowledge, this is the first report to explore the anatomical influencing factors and midterm clinical outcomes of PAI following self-expandable TAVR. The Venus-A valve was chosen, and the FEops HEARTguide simulation was used to visually exhibit interference. Additionally, the clinical and haemodynamic outcome parameters were compared between the PAI and non-PAI groups. The results showed that PAI was associated with a shorter mitral-aortic annulus distance, larger prosthesis valve size, deeper implantation, and less calcification of the non-coronary cusp. Moreover, it was associated with a higher rate of moderate or severe perivalvular leakage before discharge, but haemodynamic parameters and the incidence of adverse events at the 30-day and 12-month follow-ups were not affected.

Being in close contact with the left fibrous trigone (2,17), the AML is prone to interference with the prosthetic valve. In fact, our previous study found that in patients with pure native aortic valve regurgitation, 14 of 61 (23.0%) patients who received Venus-A prosthesis implantation had significant PAI following TAVR (18). To some extent, PAI should be considered a common complication (7,19,20), but it has rarely been described and is not covered by the VARC-3 criteria (16). In other words, the lack of consistent definitions and measurement standards in daily clinical practice has hindered the reporting of PAI. This report attempts to draw attention to this neglected clinical complication, which has been excluded from the standardised outcome reports of patients with TAVR. Given that there is currently no standardised definition, in this study, we first propose a diagnostic criterion for PAI that includes both functional and morphological assessments. To precisely describe their interaction visually, we used the FEops HEARTguide based on DSCT for computer simulation. The FEops HEARTguide has been previously reported to accurately predict the device-anatomy interaction between a self-expandable transcatheter device model and the surrounding anatomical structures (9-11), even in Chinese patients implanted with Venus-A valves (11,21,22). However, previous studies have mainly described the prosthesis frame morphology and complications such as conduction disturbance and PVL after valve implantation. In the present study, we first revealed the interaction between the prosthesis frame and the AML using the FEops HEARTguide. However, because of the limited accuracy of the FEops HEARTguide in patients who underwent valve-in-valve TAVR, this study excluded those subjects, while most valve-in-valve cases were supposed to have PAI in view of their deep implantation. In the future, improvement of the FEops HEARTguide or other technologies, such as 3D printing, may help in the evaluation of these patients.

As mentioned previously, deep implantation of a long-frame prosthesis is a significant risk factor for PAI. However, in a recently published case report, AML perforation occurred immediately after balloon predilatation due to folded leaflet calcifications distributed at the level of the non-coronary sinus toward the medial aspect of the mitral-aortic curtain (23). The authors considered that the distribution of bulky calcifications could also play a role in AML injury during TAVR procedure (23). In the present study, we further recognised several other potential risk factors, such as shorter mitral-aortic annulus distance, use of a larger prosthesis valve, and less calcification of the non-coronary cusp. The distance of the mitral aortic annulus varies across patients, and in those with prosthetic mitral valves (PMVs) undergoing TAVR, PMV-to-aortic annulus distances of <7 mm are independent risk factors for valve embolization (24). However, we could not provide a threshold value of the mitral-aortic annulus distance for risk stratification given the small sample size in this descriptive and exploratory study. Theoretically, the angle between the aortic/mitral planes should also influence their interference, but we failed to detect a difference in the mitral-aortic annulus angle between the PAI and non-PAI groups (131.4° vs 126.2°, *P*=0.333), which may be partially explained by the limited sample size. A large prosthesis was also supposed to shorten the distance between the AML and the implanted valve frame owing to its wide bottom in the LVOT. Moreover, we noticed that the difference of calcification volume at NCC was statistically significant between the groups, the reason remains speculative, but a possible explanation is that the distribution of eccentric calcification in the aortic valve facilitate deeper implantation (25).

Deep implantation may explain post-procedural PVL and new-onset left bundle branch block, but the adverse impact of PAI on 30-day and 12-month clinical prognoses was not detected in the present study, partially because of the small sample size and relatively short follow-up duration. However, we noticed that the PAI group showed a higher rate of heart failure at the 30-day and 12-month follow-ups (4/9 vs 1/11), which may be caused by a higher rate of PVL in the PAI group; the higher rate of concomitant atrial fibrillation at baseline (5/9 vs 1/11) may also worsen cardiac function. A previous study reported five cases of post-TAVR mitral valve stenosis due to PAI, with a mean trans-mitral gradient ranging from 7 mmHg to13 mmHg, three of which were treated with a conservative approach, and two others received urgent surgery (4). In the present study, the trans-mitral gradient remained at <5 mmHg in all PAI cases. This may be explained by the exclusion of valve-in-valve cases, in which patients were supposed to have a more severe impact on AML motion.

Nevertheless, the present report calls attention to this common complication regarding the devastating consequences of potential AML perforation, delayed mitral stenosis, and infective endocarditis (4-7). Furthermore, this study also emphasises the need for meticulous patient selection and strategy decision in those with a large annulus, short mitral-aortic annulus distance, and less calcification of the non-coronary cusp. To reduce the risk of PAI, a resheathable or short-frame device may be considered. Patients who may benefit from concomitant percutaneous mitral valve repair should also be carefully evaluated before developing final treatment strategies.

### 4.1 Limitations

The present study had several limitations. First, given the relatively small sample size and retrospective observational design, formal statistical analysis was not performed. Therefore, caution should be exercised when drawing firm conclusions owing to unmeasured confounders. Second, only the first-generation Venus-A valve was used in this study, and its applicability to other devices requires confirmation. However, the Venus-A valve is morphologically similar to the Medtronic CoreValve, and our findings may provide information to those who undergo CoreValve device implantation. Second, the HEARTguide is not generally used in clinical practice; simpler detection methods for PAI are needed. Third, other factors, such as extensive calcification of the aorto-mitral continuity or mitral ring, may also increase the risk of PAI and new-onset mitral valve stenosis, while no patients had severe calcification of the aorto-mitral continuity or mitral ring, and this factor was not included in the analysis. Other limitations included patient selection bias, short follow-up duration, and lack of an independent core laboratory or adjudication of clinical events.

## 5 Conclusions

PAI is associated with a shorter mitral-aortic annulus distance, larger prosthesis usage, deeper implantation, and less calcification of the non-coronary cusp. However, further studies are required to explore its long-term clinical impact.

## Data Availability

The raw/processed data required to reproduce these findings cannot be shared at this time as the data also forms part of an ongoing study.

## 6 Captions

**Supplementary Figure 1**. Description of half one cell. a) Red arrow indicates one cell, yellow arrow indicates half cell. b) Interference less than half cell.

## 8 Author contribution statement

YW and TL contributed equally to study design, data acquisition, statistical analysis, and drafted the manuscript. JJ approved the submission of the final version. YZ, PL, DQ and SY contributed greatly to data collection and the revision of the manuscript. ND, GR and SC contributed greatly to computer simulation. All authors contributed to the article and approved the submitted version.

## 11 Acknowledgments

We would like to thank Editage (www.editage.cn) for English language editing.

